# COVID-19 spreading: a recursive model

**DOI:** 10.1101/2020.04.23.20076562

**Authors:** Sergey O. Ilyin

## Abstract

This communication describes a recursive mathematical model of the spreading COVID-19 infection, which allows estimating the effectiveness of quarantine measures. This model takes into account the contagiousness of infected people during the incubation period of the disease and the conditionally non-contagiousness of sick people due to their isolation. The model was used to analyze the situation in eight countries and to find the viral transmissibility, which made it possible to give a brief prediction of the COVID-19 spreading.

---

The first mathematical models to predict the development of infectious diseases appeared in the early XX century.^1,2^ In 1927, Kermack and McKendrick proposed the use of differential equations for calculations, dividing the human population into people susceptible to disease (S) and those who had already recovered (R).^3^ The susceptible persons became infected (I) at some rate of transmission and then recovered at a different rate. Their model became known by the acronym SIR, which means that the model simultaneously calculates the number of susceptible, infected and recovered persons. This model served as a basis for the development of subsequent models by modifying the equations and adding to the calculation of other persons not belonging to the three specified basic categories, which allowed taking into account the features of particular diseases. Models have been created that consider the possibility of re-infection (SIS model)^4^ and death (SIRD),^5^ the existence of an incubation period (SEIR),^6^ temporary immunity of infants (MSIR)^7^ etc.

When a new infection appears, neither the set of population categories to be considered in the model nor the rate of transition of people from one category to another is known. Current information about the features of the novel coronavirus infection COVID-19 and the manner in which people perceive it and act should serve as a basis for building a model to describe the spread of this virus. These features are as follows: firstly, it is the presence of a long incubation period, during which the infected persons is contagious to others, and secondly, it is the isolation of discovered infected persons, which as a result become conditionally non-contagious. The combination of these two factors makes the new coronavirus infection unique. Usually the opposite is true: infected people are not dangerous to others during the incubation period and become contagious after its expiry. For this reason, a new model that considers these circumstances is needed to predict the COVID-19 spreading. Unfortunately, the duration of the immunity produced after recovery from novel coronavirus is currently unknown. In addition, there is also too little information to calculate correctly the rate of recovery of patients: a small percentage of the population recovers just a week after infection, while the majority of people are sick for a long time. Therefore, the proposed model cannot be final, but it is necessary for forecasting and management decisions.

The COVID-19 spreading model is based on a set of parameters whose values are unique for each country due to differences in population density and humans’ behavior, date of virus penetration and government actions. The set includes the following parameters:

*d*_0_ is the date of the initiation of the epidemic; it is not the date of detection of the first infected person but the date of appearance of the first undetected (or detected too late) one;
*d*_1_, *d*_2_, *d*_3_ are dates of change in the behavior of the population, e.g., due to the awareness of the reality of what is happening, the introduction of quarantine and its tightening;
*t*_D_ – is the average time from infecting to isolating the infected person, which is equal to the incubation period that I assume to be six days (from 5.2 to 6.4 days according to different sources);^8,9^ theoretically, this parameter can be reduced by total testing of the entire population, but it is feasible only for small communities;
*R*_0_, *R*_1_, *R*_2_, *R*_3_ – are the viral transmissibilities that are equal to the average number of people who will be infected by one person before its isolation and depend on the behavior of the population at different stages of the epidemic; when *R* is less than 1.0 the epidemic fades, and vice versa;
*r*_0_, *r*_1_, *r*_2_, *r*_3_ – are the reduced transmissibilities that are equal to the average number of people who will be infected by one person per day: *r* = *R*/*t*_D_; to suppress the COVID-19 spreading, *r* should be less than 0.167.

The evaluation of the virus spreading is based on the calculation of the following data:

*N*_D_(*d*_i_) is the number of detected infected persons on *d_i_* date, which equals the total number of infected persons six days earlier:

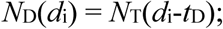

*N*_T_(*d*_i_) is the total number of infected persons on *d*_i_ date, which is the sum of the total number of infected persons the day before and the number of new infected persons that, in turns, is equal to the product of the reduced transmissibility and the number of active infected persons the day before (taking into account that those who have been previously infected cannot re-infected):

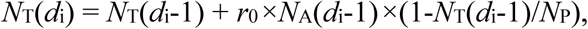

where *N*_P_ is the total population;

*N*_A_(*d*_i_) is the total number of active (undetected) infected persons on *di* date, which equals the difference between the total number of infected persons and the number of detected infected persons on the same day:

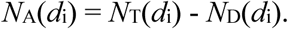

At the start of the epidemic (*d*_0_ date), *N*_A_(*d*_0_)=1, *N*_T_(*d*_0_)=1, and *N*_D_(*d*_0_)=0.

Thus, in order to calculate the virus spreading dynamics it is necessary to know the values of only two parameters – *d*_0_ and *r*_0_. In case of changing the behavior of the population from the date *d*_1_, parameter *r*_0_ changes its value from this date to become *r*_1_. If the behavior changes again, a pair of *d*_2_ and *r*_2_ will appear, etc.

It is more difficult to model human losses correctly. Two more parameters appear:

*L* is the apparent lethality rate that is equal to the ratio of the number of deaths to the sum of those who died and recovered;
*t*_L_ is the average time from infection to death.

These two parameters depend on the efficacy of treatment and may vary as physicians gain experience and as hospitals overflow. Number of deaths on *d*_i_ date equals total number of people infected *t*_L_ days earlier multiplied by the lethality rate:

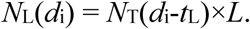

Due to the presence in the equation of two parameters (*t*_L_ and *L*) that have the same effect on the resulting value, the precision of their evaluation is lower than that of transmissibility. It should be understood that the less asymptomatic and mild cases of the disease have been detected the more the lethality rate is overestimated. The average time from infection to death was found to be about 8 days and this duration will be used to make calculations for all the countries.

The situation with predicting the number of recovered persons is even worse due to the appearance of an even greater number of independent parameters:

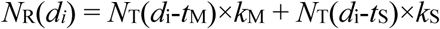

where *k*_M_, *k*_S_, *t*_M_, and *t*_S_ are the shares of mild and serious ill patients as well as the times from their infection to healing respectively; *k*_M_+*k*_S_+*L*=1.

The model equations are presented in the discrete form (instead of differential one) thanks to what for carrying out of calculations the given model is easy for reproduction in any spreadsheet editor. At first glance, it seems that the model does not take into account the existence of asymptomatic carriers of infection, but this is not true: since the share of asymptomatic carriers in the population does not change over time, their presence is taken into account implicitly by the value of the transmissibility. This model can be denoted by the SILRD abbreviation, which means that it takes into account Susceptible, Infected, isoLated, Recovered, and Dead persons.

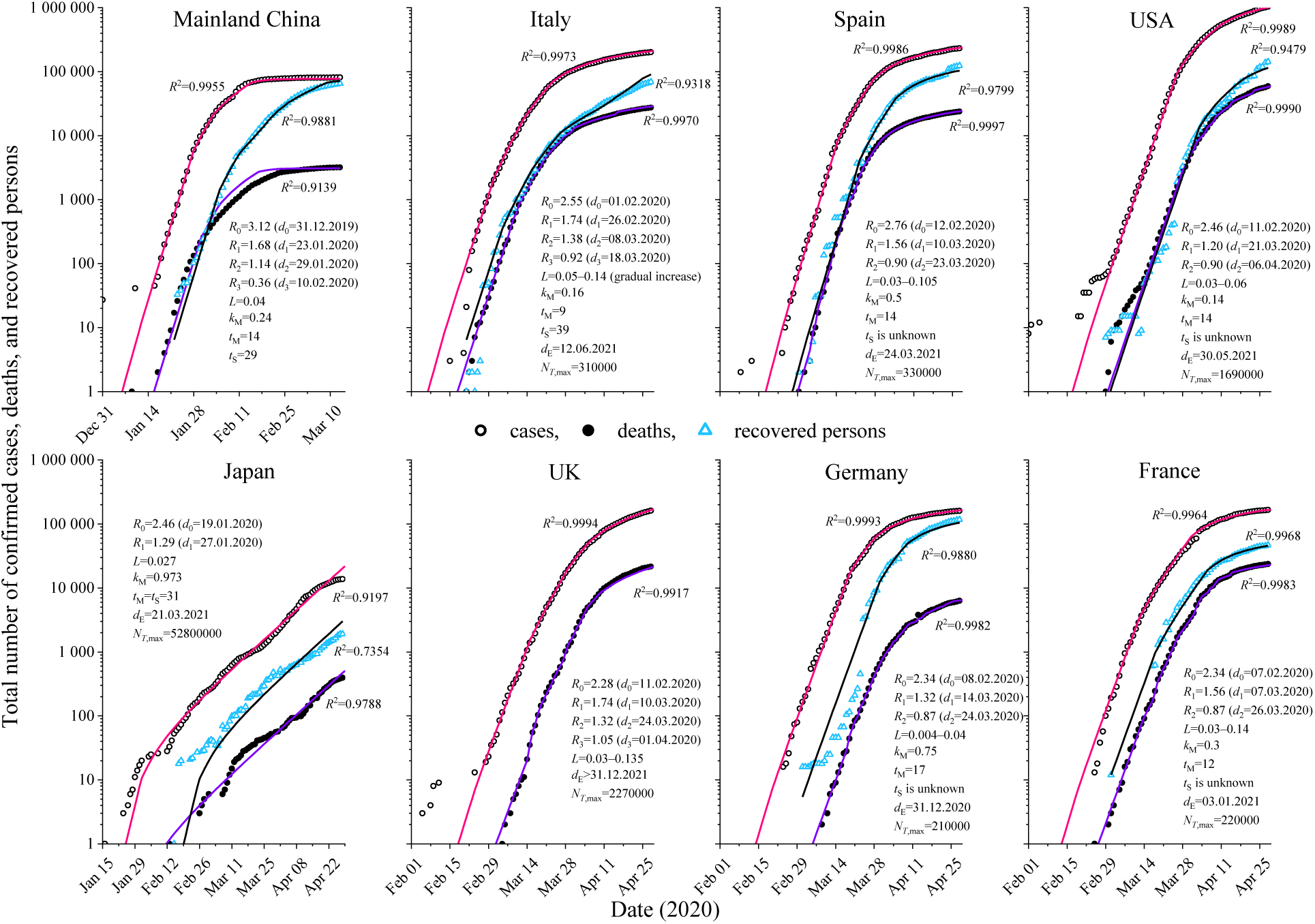
Figure. Time dependences of the total cases, deaths, and recovered from COVID-19. Dots show real data, while lines represent the result of calculations using the model.

Based on historical data on disease development in eight countries (People’s Republic of China, Italy, Spain, USA, UK, Japan, France and Germany), the model was tested and most of its parameters were found (Figure). For all the countries at the start of the epidemic, the viral transmissibility was between 2.28 and 3.12. The greatest transmissibility was found in China, where one person infected three, probably because of higher population density. The introduction and progressive strengthening of quarantine measures resulted in a decrease in the viral transmissibility, which was noticeable six days later in the decline in the rate of new cases. All the countries have introduced quarantine measures gradually. The initial restrictions reduced the viral transmissibility to 1.20–1.74, which was not enough (it was necessary to achieve transmissibility of less than 1.0) and the virus continued to spread with acceleration. As a result, all the countries, with the exception of Japan, went for tighter measures, thus reducing the transmissibility to 0.87–1.14. Japan has focused on the timely detection and isolation of the infected persons. It can be concluded that this strategy does not work: the curve of the total number of cases for Japan alternately slows down and then accelerates again. This is the result of the fact that Japan has been successfully isolating most of the infected persons, but a few people remain non-isolated and they start the epidemic again. By contrast, the PRC further strengthened the measures, which resulted in a reduction of the transmissibility to 0.36 and a quick win over the epidemic (in 6 weeks according to the model).

To date, all the European countries (except the United Kingdom) have only managed to reduce the transmissibility slightly below 1.0. From a practical point of view, this means that the number of people falling ill on a daily basis in these countries is gradually decreasing, but so slowly that the end of the epidemic in them will not be sooner, at best, than the end of this year. At the same time, it should be understood that any alleviation of quarantine measures would lead to increased transmissibility and a resumption of accelerated virus spreading. To prevent this from happening after the quarantine restrictions have been removed, the viral transmissibility must remain below 1.0. By way of example, the original transmissibility was 2.55 in the case of Italy, therefore it is necessary that 61% of the Italian population be either infected and then recovered (provided the immunity produced is durable and strong) or vaccinated against coronavirus so that when quarantine is abolished, the transmissibility remains less than 1.0. At present, 0.33% of the Italian population has been infected according to official statistics. Statistics may not take into account asymptomatic and mild cases of the disease, number of which may be 4-50 times more (for the time being only by rumor) than that of the officially recorded cases. Even if this is true, the percentage of infected and recovered persons is still significantly lower than necessary: removal of quarantine will inevitably lead to a return of the growth rate of the number of infected people to almost their original level. In other words, rapid vaccine development is vital to defeating the disease in the near future.

Thus, the model allow making a forecast of the situation development and the conclusion about the effectiveness of quarantine measures. By way of example, determine the current number of active infected persons (*N*_A_(*d*_i_)), the approximate date of isolation of the last infected person (*d*_E_), and the number of people that could eventually be infected under the current quarantine (*N*_T_,_max_). According to the calculations, the efforts made by many European countries, the USA and Japan to stop the spread of the COVID-19 infection are not as effective as those implemented previously in the People’s Republic of China. Most countries were able to achieve a daily reduction in the number of infected people, but even in these cases, the viral transmissibility remains high enough, which does not allow to defeat the epidemic within a reasonable time. At the same time, suppressing the epidemic, albeit slowly, allows time for vaccine development.

## Data Availability

All data is available on request.

